# Gestational age and infection are greater predictors of placental histopathology than maternal prepregnancy BMI

**DOI:** 10.1101/2021.12.21.21268133

**Authors:** Eleanor Duffley, David Grynspan, Hailey Scott, Anthea Lafrenière, Cherley Borba Vieira de Andrade, Enrrico Bloise, Kristin L Connor

## Abstract

The placenta undergoes morphological and functional adaptions to adverse exposures during pregnancy. The effects of suboptimal maternal body mass index (BMI), preterm birth, and infection on placental histopathological phenotypes remain unclear, despite the association between these conditions and poor offspring outcomes. We hypothesized that suboptimal maternal prepregnancy BMI and preterm birth (with and without infection) would associate with altered placental maturity and morphometry, and that altered placental maturity would associate with poor birth outcomes. Clinical data and human placentae were collected from 96 pregnancies where mothers were underweight, normal weight, overweight, or obese, without other major complications. Placental histopathological characteristics were scored with an anatomical pathologist. Associations between maternal BMI, placental pathology (immaturity and hypermaturity), placental morphometry, and infant outcomes were investigated at term and preterm, with and without infection. Fetal vascular endothelium volumetric proportion was decreased, whereas syncytial knot volumetric proportion was increased, in placentae from preterm pregnancies with chorioamnionitis compared to term placentae. At term and preterm, pregnancies with overweight and obesity had a high percentage increase in proportion of immature placentae compared to normal weight. Placental maturity did not associate with infant birth outcomes. We observed placental hypermaturity and altered placental morphometry among preterm pregnancies with chorioamnionitis, suggestive of altered placental development, which may inform about pregnancies susceptible to preterm birth and infection. Our data increase our understanding of how common metabolic exposures and preterm birth, in the absence of other comorbidities or perinatal events, potentially contribute to poor pregnancy outcomes and the programming of offspring development.

## Introduction

Maternal underweight and obesity are global health burdens and there has been a substantial increase in the prevalence of these conditions among women of reproductive age worldwide[1,2]. Mothers who are underweight or have obesity are at increased risk of delivering preterm[3], which associates with neurodevelopmental disorders and cardiometabolic diseases later in life[4-6]. The mechanisms that drive the relationship between suboptimal maternal body mass index (BMI) and adverse offspring outcomes in preterm and term pregnancies remain poorly understood, in part because cases are often confounded by multiple comorbidities and adverse perinatal events, making it difficult to disentangle the effects of specific exposures on fetoplacental development.

The placenta responds to cues in the pregnancy environment through morphological and functional changes in an effort to maintain proper fetal growth and development[7]. For example, delayed maturation of the placenta has been observed in response to increasing maternal BMI[7]. This altered placental maturity may result in poor gas and nutrient exchange at the maternal-fetal interface and, subsequently, suboptimal infant outcomes[8,9]. For example, delayed placental maturation, including the persistent thickness of vasculosyncytial membranes, forfeits optimal gas exchange and has been associated with placental insufficiency[10] and fetal macrosomia[11,12]. While the effects of maternal undernutrition on placental maturity are less studied, animal models of undernutrition have shown evidence of abnormal placental vascularization[13], which may have functional consequences. Histomorphology of the placenta ultimately determines placental function, and histological markers of placental maturity and morphometry are thus clinically useful and may reveal mechanisms underlying poor offspring outcomes in the context of suboptimal maternal BMI. Yet, the limited evidence on the effect of suboptimal maternal BMI on placental maturity and morphometry stems predominantly from complicated pregnancies, while the effects of suboptimal maternal BMI alone on placental histomorphology remain unclear.

Independent of maternal BMI, morphofunctional changes in the placenta also occur in response to increasing maternal-fetal exchange demands throughout fetal development[14]. Compared to term, preterm placentae have distinct gross and microscopic characteristics, including decreased presence of syncytial knots, thickening of the syncytiotrophoblast[15,16], and increased placental vascular lesions and evidence of malperfusion[17]; pathologies that have functional consequences for the placenta. For example, accelerated maturation of the placenta has been observed in preterm pregnancies as an attempted compensatory adaption[18], yet it associates with adverse fetoplacental outcomes, including relative placental insufficiency and late onset intrauterine growth restriction (IUGR)[19]. Moreover, chorioamnionitis due to infection of the fetal membranes results in an inflammatory cascade within fetal membranes causing premature rupture of membranes (PROM) and preterm birth (PPROM), which can further contribute to poor fetoplacental outcomes[20]. Collectively, placental (mal)adaption in response to suboptimal maternal metabolic status, infection, and/or preterm birth, may have negative effects on placental function, and thus, offspring growth and development[9]. However, the effects of suboptimal maternal BMI on placental pathological and morphometric phenotypes in preterm and term pregnancies without obstetric complications or comorbidities have been poorly quantified. This limits our understanding of how common metabolic exposures influence placental development at term and preterm and potentially contribute to the programming of offspring development.

To address this gap, we assessed the effects of suboptimal maternal prepregnancy BMI, without other major comorbidities, on histopathological indicators of placental maturity and morphometry in preterm (with and without chorioamnionitis) and term pregnancies. We also investigated the effect of gestational age and infection status on placental maturity and morphometry inclusive of maternal BMI, as gestational age and infection independently associate with altered placental histopathology. Lastly, we explored whether altered placental maturity was associated with suboptimal infant anthropometry and Apgar scores at birth. We hypothesized that maternal underweight (UW), overweight (OW), and obesity (OB) would associate with suboptimal placental maturity and morphometry, and that altered placental maturity would associate with poor infant birth anthropometry and Apgar scores. By characterizing the placental pathological and morphological phenotypes of pregnancies complicated by suboptimal maternal metabolic status, preterm birth, and infection, our work may uncover mechanisms that can explain poor offspring development in these pregnancies, and placental-specific histological markers that could predict altered postnatal health trajectories.

## Methods

### Study population

This study was approved by the Mount Sinai Hospital Research Ethics Board (17-0186-E) and the Carleton University Research Ethics Board (106932). Clinical data and human placentae from 96 pregnancies were collected through the Research Centre for Women’s and Infants’ Health (RCWIH) BioBank at Mount Sinai Hospital, Toronto. Inclusion criteria were singleton pregnancies and live birth with no known fetal anomalies. Exclusion criteria were gestational diabetes mellitus, hypertension (including pregnancy induced hypertension), HELLP syndrome, lupus, antiphospholipid antibody syndrome, Crohn’s disease, ulcerative colitis, colitis, Guillain-Barré syndrome, sexually transmitted infections, gastritis, urinary tract infections, smokers, documented recreational drug use during pregnancy, pelvic inflammatory disease, and *in vitro* fertilization. Women were categorized as having delivered preterm with chorioamnionitis (PTC, n= 29), preterm without chorioamnionitis (PT, n= 31), or at term (T, n= 36; there were no term pregnancies with chorioamnionitis), and were further classified as underweight (UW, n=21), normal weight (NW, n=24), overweight (OW, n=27) or having obesity (OB, n=24). Gestational age was calculated based on the last menstrual period to the nearest week. Among 60 preterm pregnancies, 12 women were classified as UW by prepregnancy BMI, 17 as NW, 18 as OW, and 13 as OB, and among 36 term pregnancies, nine were classified as UW, seven NW, nine OW, and 11 OB.

### Maternal and infant characteristic data collection

The primary exposure of interest was maternal prepregnancy BMI classified according to the World Health Organization and American College of Obstetricians and Gynecologists guidelines [21] with one exception: due to a low prevalence of women considered underweight in the study region, a prepregnancy BMI of less than 19 kg/m^2^ was considered to be underweight (UW). Maternal underweight, overweight (≥25–29.9 kg/m^2^), and obesity (≥30.0 kg/m^2^) were compared to normal weight controls (19–24.9 kg/m^2^). Maternal prepregnancy BMI was considered as both a continuous and categorical exposure variable. A retrospective medical chart review was conducted to extract antenatal and birth data, including maternal characteristics (prepregnancy height and weight, age at conception, ethnicity, gestational weight gain). Infant data included gestational age, infant sex, Apgar scores (at one, five, and ten minutes), and newborn anthropometry (including birthweight, a secondary outcome). Standardized birthweight by infant sex and gestational age were calculated based on singleton data reported by Kramer et al[22].

### Placental collection and processing

Placentae were collected by the RCWIH BioBank and biopsies were processed for histology. Formalin-fixed, paraffin embedded placental biopsies were sectioned (6 μm) and stained with haematoxylin (Gill’s Number 1, Sigma-Aldrich) and eosin (Eosin Y-Solution, Sigma-Aldrich) (H&E) according to standard protocols. The primary outcomes of interest were placental pathologies, specifically microscopic placental pathologies related to morphometry and those indicative of placental maturity.

### Placental morphometry

Placental morphometry analysis was undertaken using methodology previously described in the literature with specific adaptations, outlined below[23]. Image acquisition and analysis were performed on H&E-stained sections using an Aperio AT2 microscope (Leica Biosystems, IL, EUA), coupled with a computer using the software ImageScope x64. The histomorphological analysis was undertaken by an experienced examiner, blinded to exposure groups, using the Fiji ImageJ 1.0 program (ImageJ, WI, USA). Relative volume estimates of placental histological components (syncytiotrophoblast, syncytial knots, cytotrophoblasts, connective tissue, and fetal vascular endothelium) were quantified by superimposing placental histological photomicrographs with a grid of equidistant points (measuring 25 μm distance between two points). Previous studies using morphometric analysis in human placentae recorded 190-1000 points (grid intercepts) to evaluate the volumetric proportion of each placental histological component[23-25]. In this study, we first assessed placentae by recording 1500 points overlapping with each of the histological components for the first 39 placentae. For the remaining 48 placentae, in order to optimize the recordings, we reduced the number of points recorded to 600 points overlapping with each of the histological components, while still exceeding the number of points typically assessed[25]. We analysed volumetric proportions of each histological component across BMI or gestational age groups for placentae from 1500 and 600 point morphometric analyses separately. There were no differences in volumetric proportions of histological components between the placentae assessed using the 1500 point approach and the 600 point approach when comparing outcomes across maternal BMI or gestational age groups (data not shown). Thus, placentae assessed using 1500 and 600 points were pooled for statistical analyses. The total average area of evaluated histological sections per placenta was 393,173.22 μm^2^, and there were no differences in median area (μm^2^) assessed across maternal BMI groups (UW: 324043 [273407, 587270]; NW: 505332 [259336, 557864]; OW: 276020 [257202, 505674]; OB: 502106 [270320, 532097]). The volumetric proportion (VP) of each histological component was calculated as VP = NP ×100/600 for placentae for which 600 points were recorded, and as VP = NP ×100/1500 for placentae for which 1500 points were recorded, where NP = number of equivalent points on each histological component[23,26,27].

### Placental maturity

Histopathological characteristics were scored on H&E-stained sections by an anatomical pathologist following the Amsterdam criteria[28] to assess for placental maturity relative to gestational age. Characteristics of immaturity and hypermaturity were scored as either 0 (absent) or 1 (present). Characteristics of immaturity were: 1. villous immaturity [monotonous villi (≥10 villi?) with centrally placed capillaries] and 2. stromal immaturity [decreased vasculosyncytial membranes resembling villi in early pregnancy present in at least 30% of full thickness section]. Characteristics of hypermaturity were: 1. hypercapillarisation [chorangiosis defined as >10 terminal villi with ≥10 capillaries], 2. distal villous hypoplasia [reduced size of intermediate villi with dispersed terminal villi and reduced number that appear thin and elongated, widening of intervillous space; adjusted for gestational age; involving at least 30% of a full thickness slide], and 3. accelerated maturity [the presence of term-appearing and/or hypermature villi for gestational age, not in areas adjacent to infarction]. No placentae had both immature and hypermature characteristics. Placentae with presence of one or more immaturity characteristic but absent of all hypermaturity characteristics were deemed ‘immature’ for gestational age. Placentae with presence of one or more hypermaturity characteristic but absent of all immaturity characteristics were deemed ‘hypermature’ for gestational age. Placentae absent of all immaturity and hypermaturity characteristics were deemed ‘normal’ for gestational age.

### Statistical analyses

#### Univariate analyses

The primary exposure of interest was maternal prepregnancy BMI, specifically maternal UW, OW, and OB compared to NW controls. As gestational age and infection status can independently affect outcomes, we also assessed these variables as secondary exposures. The primary outcomes of interest were placental maturity (immature, normal, hypermature) and placental morphometry (syncytiotrophoblast, syncytial knots, cytotrophoblasts, connective tissue, and fetal vascular endothelium). Data were stratified by term (37 – 42.2 weeks gestation) and preterm (<37 weeks gestation) to assess the relationships between maternal prepregnancy BMI and outcome variables in preterm and term pregnancies separately. Associations between maternal prepregnancy BMI groups or preterm/term group and continuous outcomes (placental morphometry measures) were tested using one-way ANOVA or Kruskal– Wallis test with Tukey’s *post hoc* or Steel–Dwass *post hoc*. Likelihood Ratio Chi Square tests were used to evaluate the associations between maternal BMI groups or preterm/term group and categorical outcome variables (placental maturity outcomes). To explore sex differences in placental maturity and morphometry in response to both BMI and gestational age, we also stratified data by fetal sex to analyze outcomes in males and females separately. Data were analysed using JMP statistical software (version 14.2). Data are presented as median (interquartile range; non-parametric data), mean, and standard deviation (parametric data) or frequency (percentage; categorical variables). Statistical significance was defined as p<0.05.

#### Multivariable analyses

Multivariable regression analyses were conducted to assess the relationship between maternal prepregnancy BMI (continuous) and placental maturity and morphometry separately for preterm and term pregnancies. First, an unadjusted model was used to identify the associations between prepregnancy BMI with placental maturity in preterm and term pregnancies (model A). An adjusted ordinal logistic regression model was then used to determine the associations between maternal BMI and placental maturity (model B) adjusted for fetal sex (male/female), and gestational weight gain (continuous) for term pregnancies, and also adjusting for chorioamnionitis (yes/no) and gestational age (continuous) for preterm pregnancies. Data are presented as β (or adjusted β [aβ]) and 95% confidence intervals, and p value from Likelihood Ratio Chi Square test. Thirdly, an unadjusted model was used to identify the associations between prepregnancy BMI and placental morphometry data (model C). An adjusted standard Least Squares regression model was used to determine the associations between maternal BMI and placental morphometry data (model D; adjusted for fetal sex [male/female], and gestational weight gain [continuous] for term pregnancies and was also adjusted for chorioamnionitis [yes/no], and gestational age [continuous] for preterm pregnancies). Data are presented as β (or adjusted β [aβ]) and 95% confidence intervals, and p value from Standard Least Squares regression models.

## Results

### Maternal BMI has limited effect on placental maturity or morphometry

There was no effect of maternal prepregnancy BMI on placental anthropometry among preterm or term pregnancies (Table 1). However, at term, placental immaturity was more prevalent in OW and OB pregnancies, representing a 400% increase in proportion of placentae that were immature in OW and OB, respectively, compared to NW pregnancies. At preterm, placental immaturity was more prevalent in OW and OB preterm pregnancies, representing a 400% and 200% increase in proportion of placentae that were immature in OW and OB, respectively, compared to NW pregnancies. However, among preterm and term pregnancies, there were no statistical differences in placental maturity (immature, normal, or hypermature classifications) across maternal BMI groups (Figure 1). Placental maturity also did not differ with increasing maternal BMI among preterm pregnancies ([model A: β= 0.06 (−0.007, 0.15), p= 0.07]; [model B: aβ=0.04 (−0.04, 0.12), p= 0.33]) or term pregnancies ([model A: β= 0.05, (−0.04, 0.16), p= 0.27]; [model B: aβ=0.06 (−0.04, 0.21), p= 0.24]). When data were further stratified by fetal sex, there were no differences in placental maturity across maternal BMI groups at preterm and term in male or female placentae (Supplementary Tables 1 and 2).

**Table 1.** Infant characteristics by prepregnancy BMI in preterm and term pregnancies.

|  | Prepregnancy BMI |  |  |  |  |
| --- | --- | --- | --- | --- | --- |
|  | UW | NW | OW | OB | p-value |
| Infant characteristics |  |  |  |  |  |
| Preterm pregnancies | (n=12) | (n=17) | (n=18) | (n=13) |  |
| Sex (n [%]) |  |  |  |  | 0.47 |
| Female | 5 (41.7) | 5 (29.4) | 10 (55.6) | 5 (38.5) |  |
| Male | 7 (58.3) | 12 (70.59) | 8 (44.4) | 8 (61.5) |  |
| Placental weight (g) | 410 (350, 490) | 400 (330, 530) | 448 (323, 690) | 480 (388, 670) | 0.21 |
| Z-birthweight | -0.34 ± 0.94<sup>A</sup> | -0.02 ± 0.71<sup>AB</sup> | 0.20 ± 0.65<sup>AB</sup> | 0.59 ± 1.09<sup>B</sup> | **0.05** |
| Birthweight (g) : placental weight (g) | 4.39 ± 1.19 | 3.96 ± 1.23 | 3.73 ± 1.19 | 3.90 ± 1.00 | 0.52 |
| Apgar score |  |  |  |  |  |
| 1 minute | 8 (6.25, 8) | 8 (4.5, 9) | 8 (5, 9) | 8.5 (3.75, 9) | 0.93 |
| 5 minutes | 8.5 (8, 9) | 9 (8, 9) | 9 (8, 9) | 9 (8, 9) | 0.87 |
| 10 minutes | 8 (8, 8) | 8 (2.75, 8.75) | 7 (7, 9) | 9 (5, 9) | 0.90 |
| Gestational age (weeks) | 32.1 (29, 34.2) | 31.3 (27, 34.7) | 31 (27.5, 34.8) | 33.3 (29.6, 35.2) | 0.81 |
| Term pregnancies | (n=9) | (n=8) | (n=8) | (n=11) |  |
| Sex (n [%]) |  |  |  |  | 0.05 |
| Female | 3 (33.3) | 2 (37.5) | 6 (66.7) | 9 (81.8) |  |
| Male | 6 (66.7) | 5 (62.5) | 3 (33.3) | 2 (18.2) |  |
| Placental weight (g) | 500 (465, 620) | 700 (563, 743) | 610 (550, 780) | 618 (570, 1000) | 0.09 |
| Z-birthweight | -0.31 (-0.78, 0.11)<sup>A</sup> | 0.02 (-0.49, 0.75)<sup>A</sup> | 0.14 (-0.27, 0.66)<sup>A</sup> | 0.62 (0.09, 2.28)<sup>A</sup> | **0.04** |
| Birthweight (g) : placental weight (g) | 6.11 ± 0.87 | 5.45 ± 1.00 | 5.46 ± 0.74 | 5.15 ± 1.35 | 0.25 |
| Apgar score (n [%]) |  |  |  |  |  |
| 1 minute | 9 (8, 9) | 9 (9, 9) | 9 (8.5, 9) | 9 (6.75, 9) | 0.47 |
| 5 minutes | 9 (9, 9) | 9 (9, 9) | 9 (9, 9) | 9 (8.75, 9) | 0.37 |
| 10 minutes | - | - | 9 | 9 (9, 9) | 1.00 |
| Gestational age (weeks) | 39.3 ± 0.52 | 39.3 ± 1.05 | 39.8 ± 1.30 | 38.7 ± 0.88 | 0.12 |

Data are means ± SD (ANOVA; normal distribution/equal variance) or median (IQR; Kruskal-Wallis/Wilcoxon test for non-parametric data with Steel-Dwass *post-hoc*). Data are n (%) (Likelihood Ratio Chi Square test) for categorical variables. \*p<0.05.

**Fig. 1.**
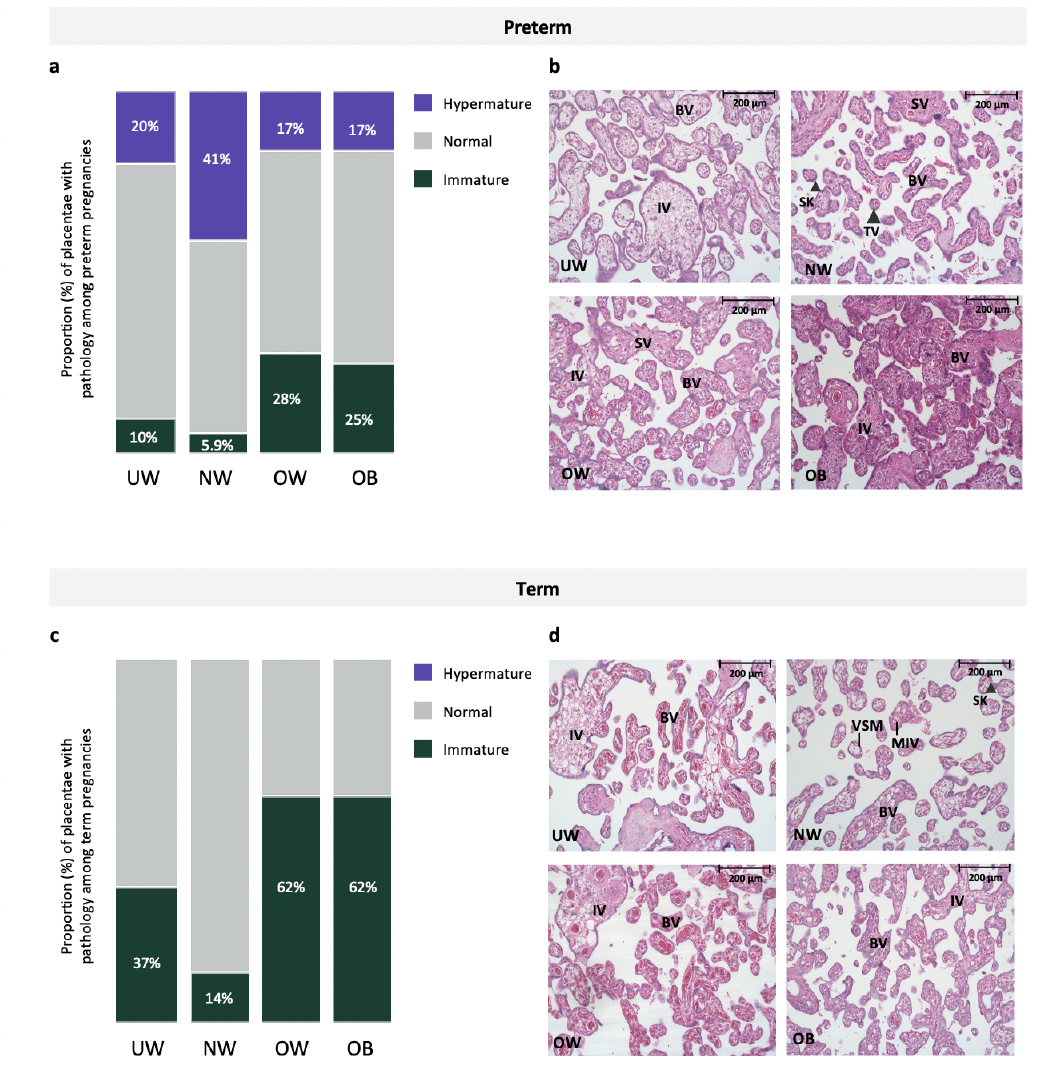
a. Proportion of placentae with immature, normal, and hypermature placentae across BMI and gestational age groups among preterm pregnancies. Placental immaturity was more prevalent in OW and OB pregnancies, representing a 400% and 200% increase in proportion of placentae that were immature in OW and OB, respectively, compared to NW pregnancies. b. Representative images from H&E-stained placentae from UW, NW, OW and OB preterm pregnancies. UW, OW, OB=immature pathology. BV=Blood vessel, IV=immature villus, TV=terminal villus, VSM=Vasculo-syncytial membrane, SK (small arrowhead)=Syncytial knot, SV=Stem villus. 20X Magnification. Scale bar = 200 μm. c. Proportion of placentae with immature, normal, and hypermature placentae across BMI and gestational age groups among term pregnancies. At term, placental immaturity was more prevalent in OW and OB pregnancies, representing a 400% increase in proportion of placentae that were immature in both OW and OB, compared to NW pregnancies. No hypermaturity was observed in term placentae. d. Representative images from H&E-stained placentae from UW, NW, OW and OB term pregnancies. UW, OW, OB=immature pathology. BV=Blood vessel, IV=immature villus, MIV=mature intermediate villus, TV (large arrowhead)=terminal villus, VSM=Vasculo-syncytial membrane, SK (small arrowhead)=Syncytial knot. 20X Magnification. Scale bar = 200 μm

Among preterm and term pregnancies, connective tissue comprised the greatest area of quantified tissue, followed by fetal vascular endothelium, syncytiotrophoblast, and syncytial knots, and there were no differences in volumetric proportion of these histologic features across maternal BMI groups (Figure 2). Placental morphometry also did not differ with increasing maternal BMI (Table 2). When data were stratified by fetal sex, there was no association between maternal BMI and volumetric proportion of any histological components at preterm and term in male or female placentae (Supplementary Tables 1 and 2).

**Table 2.** Associations between pre-pregnancy BMI and placental morphometry volumetric proportions in preterm and term pregnancies.

| Volumetric proportion of placental histological components (%) | Model C |  | Model D |  |
| --- | --- | --- | --- | --- |
| | $\beta$ (95% CI) | p-value | aβ (95% CI) | p-value |
| **Preterm (n= 60)** |  |  |  |  |
| Syncytiotrophoblast | -0.01 (-0.15, 0.12) | 0.80 | -0.05 (-0.19, 0.08) | 0.42 |
| Cytotrophoblast | 0.001 (-0.006, 0.008) | 0.74 | -0.002 (-0.008, 0.004) | 0.53 |
| Connective tissue | -0.19 (-0.48, 0.09) | 0.18 | -0.22 (-0.53, 0.08) | 0.16 |
| Fetal Vascular endothelium | 0.21 (-0.11, 0.54) | 0.19 | 0.28 (-0.05, 0.63) | 0.09 |
| Syncytial knots | -0.008 (-0.04, 0.02) | 0.60 | -0.01 (-0.04, 0.02) | 0.52 |
| **Term (n= 36)** |  |  |  |  |
| Syncytiotrophoblast | -0.11, (-0.29, 0.06) | 0.18 | -0.08 (-0.31, 0.1) | 0.42 |
| Cytotrophoblast | -0.006 (-0.02, 0.009) | 0.42 | -0.002 (-0.02, 0.02) | 0.82 |
| Connective tissue | 0.03 (-0.31, 0.38) | 0.81 | -0.07 (-0.52, 0.38) | 0.73 |
| Fetal Vascular endothelium | 0.06 (-0.32, 0.45) | 0.74 | 0.14 (-0.37, 0.66) | 0.58 |
| Syncytial knots | -0.005 (-0.03, 0.02) | 0.74 | -0.002 (-0.04, 0.03) | 0.63 |

Data are $\beta$ and 95% CI and p value from Standard Least Squares models. \* $p < 0.05$ . For preterm, model B adjusted for fetal sex, maternal gestational weight gain, chorioamnionitis status, and gestational age. For term, model B includes fetal sex and maternal gestational weight gain.

**Fig. 2.**
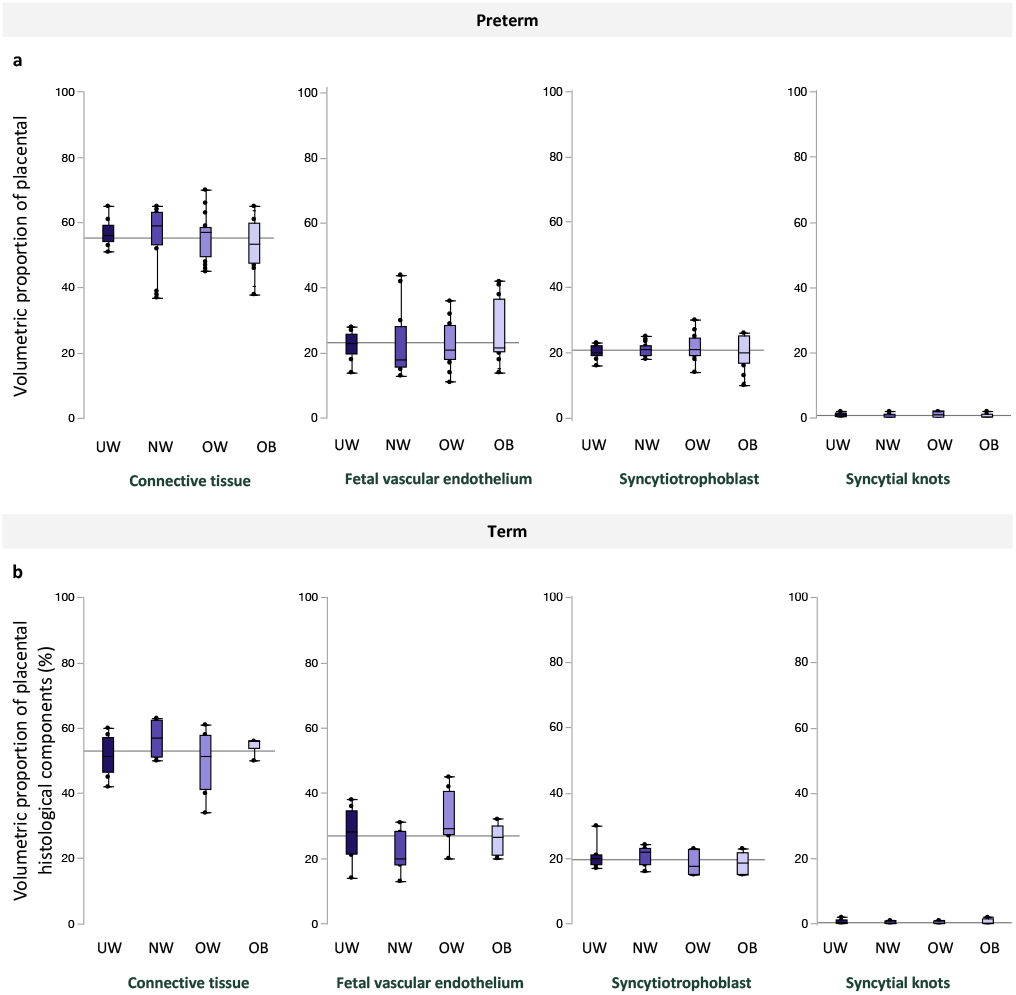
Volumetric proportion of placental histological components (%), including, from left to right, fetal vascular endothelium, connective tissue, syncytiotrophoblast, and syncytial knots across maternal BMI groups among a. preterm and b. term pregnancies. Data are quantile box plots with a horizontal line representing the mean across the whole cohort

### Maternal BMI has limited effect on birth outcomes

At birth, standardized birthweight increased with increasing maternal BMI among preterm and term infants (Table 1), although among term infants there were no differences in infant birthweight z-scores (BWZ) between BMI groups on *post hoc* analysis. There were no differences across maternal BMI groups for infant Apgar scores at 1 or 5 minutes among preterm or term infants (Table 1).

### Gestational age and infection status associate with altered placental maturity and morphometry

We found that placental weight (p<0.0001) and birthweight to placental weight ratio (p<0.0001), a proxy measure for placental efficiency, were decreased in preterm pregnancies with chorioamnionitis compared to preterm without chorioamnionitis and term pregnancies, inclusive of BMI (Table 3). Additionally, inclusive of BMI, the greatest proportion of placental hypermaturity was observed in preterm pregnancies with chorioamnionitis [immature = 1 (3.7), normal = 15 (55.6), hypermature = 11 (40.7)] compared to preterm [immature = 8 (27.6), normal= 18 (62.1)], hypermature = 3 (10.3)] and term [immature = 14 (45.2), normal = 17 (54.8), hypermature = 0 (0.00)] pregnancies (p<0.0001). Prevalence of immaturity, normal maturity, and hypermaturity did not differ across gestational age/infection groups when stratified by fetal sex (Figure 3, Supplementary Table 3).

**Table 3.** Infant characteristics in preterm pregnancies with chorioamnionitis, preterm pregnancies without chorioamnionitis, and term pregnancies.

|  | Gestational age and infection status |  |  | p-value |
| --- | --- | --- | --- | --- |
|  | Preterm Chorio<br>(n=29) | Preterm<br>(n=31) | Term<br>(n=36) |  |
| Infant characteristics |  |  |  |  |
| Sex (n [%]) |  |  |  | 0.37 |
| Female | 13 (44.8) | 12 (38.7) | 20 (55.6) |  |
| Male | 16 (55.7) | 19 (61.3) | 16 (44.4) |  |
| Placental weight (g) | 368 (303, 463)<sup>A</sup> | 500 (400, 650)<sup>B</sup> | 600 (543, 743)<sup>C</sup> | <0.0001 |
| Z-birthweight | 0.02 (-0.34, 0.33) | 0.3 (-0.71, 0.89) | 0.14 (-0.49, 0.74) | 0.85 |
| Birthweight (g) : placental weight (g) | 3.43 ± 1.04<sup>A</sup> | 4.43 ± 1.05<sup>B</sup> | 5.54 ± 1.05<sup>C</sup> | <0.0001 |
| Apgar score |  |  |  |  |
| 1 minute | 8 (4, 8)<sup>A</sup> | 8.5 (6, 9)<sup>AB</sup> | 9 (8, 9)<sup>B</sup> | 0.0003 |
| 5 minutes | 8 (6, 9)<sup>A</sup> | 9 (8, 9)<sup>A</sup> | 9 (9, 9)<sup>B</sup> | 0.0001 |
| 10 minutes | 7 (5, 8)<sup>A</sup> | 9 (8, 9)<sup>B</sup> | 9 (9, 9)<sup>B</sup> | 0.0048 |
| Gestational age (weeks) | 28.1 (26.6, 31.4)<sup>A</sup> | 34.7 (31.9, 35.6)<sup>B</sup> | 39.1 (38.6, 39.7)<sup>C</sup> | <0.0001 |

Data are means ± SD (ANOVA; normal distribution/equal variance with Tukey *post-hoc*) or median (IQR; Kruskal-Wallis/Wilcoxon test for non-parametric data with Steel-Dwass *post-hoc*). Data are n (%) (Likelihood Ratio Chi Square test) for categorical variables. \*p<0.05.

**Fig. 3.**
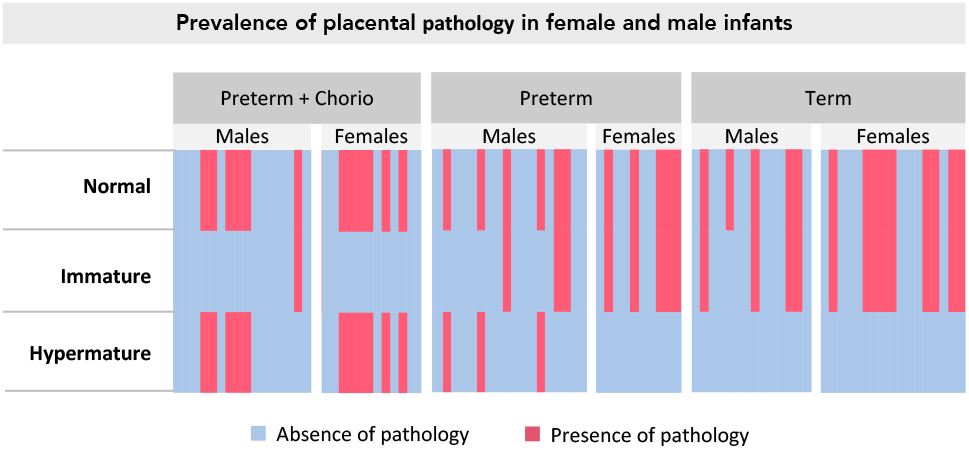
Heat map of placental maturity prevalence amongst preterm with chorioamnionitis (males n=16, females n=13), preterm (males n=19, females n=12), and term pregnancies (males n=16, females n=20) and stratified by fetal sex. Blue represents absence of pathology and red represents presence of pathology (immaturity [villous immaturity and stromal immaturity] or hypermaturity [hypercapillarisation, distal villous hypoplasia, and accelerated maturation]).

Fetal vascular endothelium volumetric proportion was decreased (p=0.04, Table 4) and syncytial knot volumetric proportion was increased (p=0.02, Table 4) in preterm pregnancies with chorioamnionitis compared to term. However, gestational age had no effect on connective tissue, syncytiotrophoblast, or cytotrophoblast volumetric proportions (Table 4). When data were stratified by fetal sex, fetal vascular endothelium volumetric proportion was decreased (p=0.03, Supplementary Table 4) and syncytial knot volumetric proportion was increased (p=0.006, Supplementary Table 4) in female placentae from preterm pregnancies with chorioamnionitis compared to female placentae from term pregnancies. Gestational age had no effect on connective tissue or syncytiotrophoblast, or cytotrophoblast volumetric proportions in female placentae. There were no differences in volumetric proportion of histologic features in male placentae across gestational age groups.

**Table 4.**
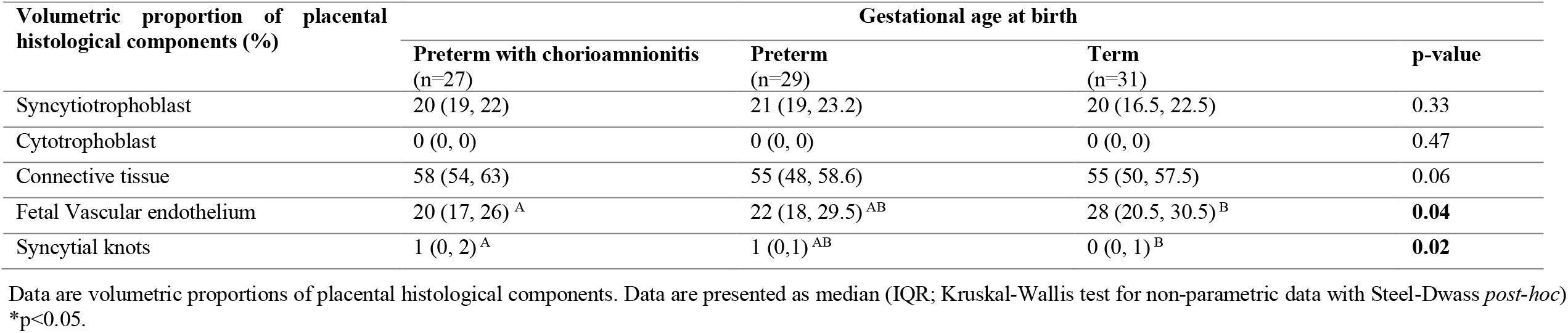
Effect of gestational age at birth and infection status on volumetric proportion of placental histological components.

| Volumetric proportion of placental histological components (%) | Gestational age at birth |  |  | p-value |
| --- | --- | --- | --- | --- |
|  | Preterm with chorioamnionitis<br>(n=27) | Preterm<br>(n=29) | Term<br>(n=31) |  |
| Syncytiotrophoblast | 20 (19, 22) | 21 (19, 23.2) | 20 (16.5, 22.5) | 0.33 |
| Cytotrophoblast | 0 (0, 0) | 0 (0, 0) | 0 (0, 0) | 0.47 |
| Connective tissue | 58 (54, 63) | 55 (48, 58.6) | 55 (50, 57.5) | 0.06 |
| Fetal Vascular endothelium | 20 (17, 26) <sup>A</sup> | 22 (18, 29.5) <sup>AB</sup> | 28 (20.5, 30.5) <sup>B</sup> | 0.04 |
| Syncytial knots | 1 (0, 2) <sup>A</sup> | 1 (0,1) <sup>AB</sup> | 0 (0, 1) <sup>B</sup> | 0.02 |

Data are volumetric proportions of placental histological components. Data are presented as median (IQR; Kruskal-Wallis test for non-parametric data with Steel-Dwass *post-hoc*) \*p<0.05.

### Preterm pregnancies with chorioamnionitis associate with decreased infant Apgar scores

Inclusive of maternal BMI, there were no differences in infant BWZ between preterm, preterm with chorioamnionitis, and term pregnancies (Table 3). However, Apgar scores at one minute (p=0.0003) were decreased in preterm pregnancies with chorioamnionitis, compared to scores in term infants, and Apgar scores at 5 minutes were also decreased in both preterm pregnancies with and without chorioamnionitis compared to term infants (p=0.0003, Table 3). Preterm pregnancies with chorioamnionitis also had the lowest median gestational age at birth, followed by preterm pregnancies without infection and term pregnancies (p<0.0001, Table 3). Placental maturity did not associate with infant BWZ or Apgar scores at 1 and 5 minutes (Figure 4) among preterm and term pregnancies.

**Fig. 4.**
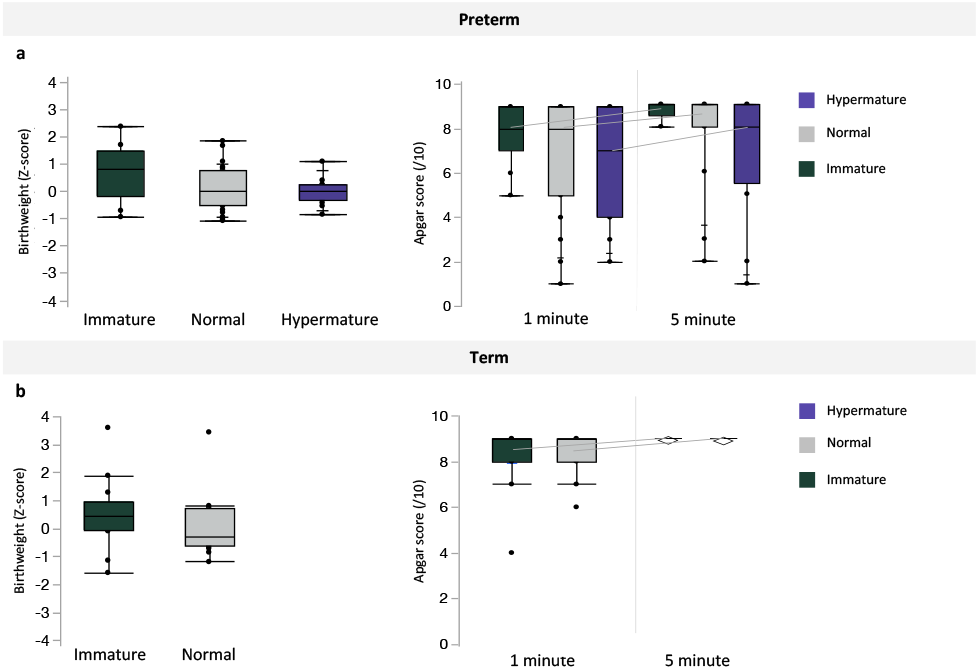
Associations between placental maturity (immature, normal, and hypermature) and birthweight z-scores or Apgar scores at 1 and 5 minutes among a. preterm and b. term pregnancies. Data are quantile box plots.

## Discussion

We examined the effect of maternal prepregnancy BMI, without other major comorbidities, on placental maturity and morphometry to quantify how suboptimal maternal metabolic states influence placental phenotypes and to better understand the mechanisms that may contribute to poor pregnancy outcomes and fetal (mal)development in these pregnancies. Reassuringly, we found no major differences in placental maturity or morphometry across maternal prepregnancy BMI groups among preterm or term pregnancies and placental maturity did not associate with infant birthweight or Apgar scores at birth. There were limited associations between maternal BMI and infant birth outcomes. We did observe an effect of gestational age and infection on placental phenotypes, where the greatest proportion of hypermature placentae were from preterm pregnancies with chorioamnionitis, compared to preterm without infection and term pregnancies. In contrast, preterm pregnancies with chorioamnionitis associated with decreased placental weight and efficiency (as correlated with infant birthweight to placental weight ratio) and infant Apgar scores, confirming that preterm birth with infection impacts placental maturity, placental function, and newborn outcomes.

Our data showed limited evidence for an effect of low or high prepregnancy BMI on placental maturity and morphometry among preterm or term pregnancies. Among both term and preterm pregnancies, we found that placental immaturity was more prevalent in OW and OB pregnancies, representing a high percentage increase in the proportion of placentae that were immature in both OW and OB pregnancies, compared to NW pregnancies, which may suggest the emergence of underlying pathology. It is well established that maternal obesity promotes a pro-inflammatory environment within gestational tissues, including elevated levels of circulating interleukin [IL]-6 during pregnancy and higher levels of placental pro-inflammatory cytokines^9,^ and associates with poor infant outcomes. Diet-induced maternal obesity also associates with increased levels of the proinflammatory cytokines tumor necrosis factor [TNF]-α and IL-8 in sheep placentae[29]. TNF-α can inhibit placental trophoblast motility and migration, indicating its potential to impact placental development^14-16^. Thus, poor maternal metabolic health is permissive of a pro-inflammatory environment that may adversely affect normal placental maturity and structure. However, in contrast with studies showing increased proportion of macroscopic and microscopic placental pathologies with increasing maternal BMI[7,30], our data show limited evidence for altered placental histopathology in pregnancies with suboptimal maternal BMI. Differences with our findings could be explained by our study design. We intentionally excluded pregnancies with comorbidities and complications that are associated with obesity and underweight and have known effects on placental development and function[12,31] so that we could more accurately gain insight into the effects of suboptimal BMI alone on placental pathology and morphometry. A systemic pro-inflammatory state is present in women with pre-eclampsia and gestational diabetes mellitus[32,33], and placental pathologies, such as villous immaturity or chorangiosis, have been reported in women with these complications[11,12,34], irrespective of maternal BMI. Hypertension and gestational diabetes mellitus are highly prevalent in mothers who are overweight or have obesity[35] and could be driving the placental histopathological changes previously reported in pregnancies complicated by suboptimal maternal BMI. Given that an estimated 30% of women with overweight or obesity have no other comorbidities[36,37], our limited histomorphological findings in placentae from otherwise uncomplicated pregnancies may be reassuring.

Gestational age and infection may also alter placental maturity and morphometry. Indeed, our placental morphometric analyses show decreased fetal vascular endothelium volumetric proportion in preterm pregnancies with chorioamnionitis compared to term pregnancies. Endothelial cell proliferation and elongation is critical for placental vascular remodeling throughout pregnancy[38], and placental endothelial cell dysfunction can contribute to the development of disorders such as placental insufficiency and pre-eclampsia[39]. Thus, decreased fetal vascular endothelium in preterm pregnancies with chorioamnionitis may suggest inadequate placental vasculature and decreased placental blood flow throughout gestation. In what may have been an attempted compensatory adaptation to decreased fetal vascular endothelium, placental blood flow, and subsequent placental hypoxia [40], preterm pregnancies with chorioamnionitis also showed a greater proportion of placental hypermaturity compared to preterm and term pregnancies (proposed mechanism depicted in Figure 5). This is consistent with previous studies suggesting that placental villous hypermaturity in idiopathic preterm pregnancies and preterm pregnancies with chorioamnionitis may be an attempt to increase nutrient exchange[41]. We also observed increased volumetric proportion of syncytial knots in placentae from preterm with chorioamnionitis compared to term placentae, consistent with a hypermature placental phenotype [42,43]. Syncytiotrophoblast shedding via increased number of syncytial knots may thus be one mechanism leading to placenta hypermaturity in preterm pregnancies with chorioamnionitis. Further, given that altered placental morphometry represents placental changes throughout gestation (and not acute response to infection), decreased fetal vascular endothelium volumetric proportion merits further investigation as a potential indicator for placentae at risk of later preterm birth and infection/chorioamnionitis.

**Fig 5.**
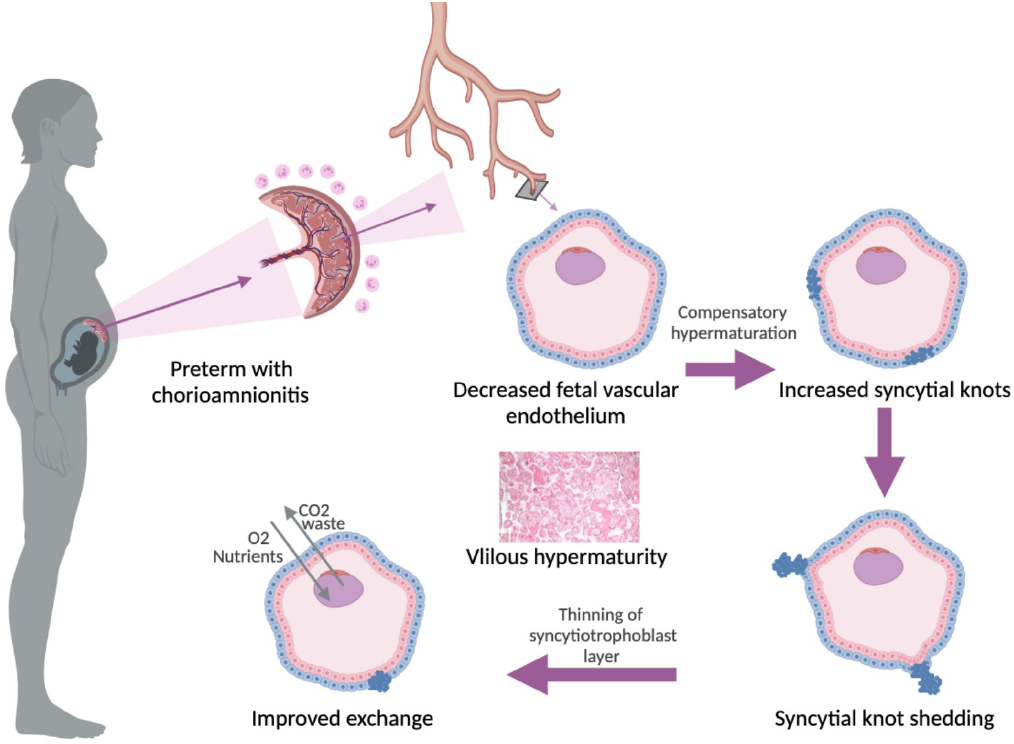
Proposed mechanism of placental villous hypermaturity in preterm pregnancies with chorioamnionitis. Decreased fetal vascular endothelium, and subsequently inadequate placental vasculature and blood flow, in preterm pregnancies with chorioamnionitis may prompt compensatory placental hypermaturation. Syncytiotrophoblast shedding via increased number of syncytial knots and subsequent thinning of the syncytiotrophoblast layer may be one mechanism leading to placental villous hypermaturity and improved exchange in preterm pregnancies with chorioamnionitis.

Optimal placental function is necessary for the delivery of nutrients, oxygen, and hormones to the developing fetus[44]. The histological markers of placental immaturity and hypermaturity are indicative of a placenta that may be structurally ill-suited to meet fetal demands[11,12,19]. However, in our cohort, placental maturity did not associate with infant outcomes at birth in preterm or term pregnancies. This is surprising, as previous studies have supported the prognostic value of placental histology, including demonstrating associations between placental maturity and infant outcomes[40,45]. Whereas previous studies linking placental maturity and infant birth outcomes were often from complicated pregnancies, our cohort purposefully lacked major maternal comorbidities apart from suboptimal maternal BMI. This suggests that altered placental maturity may only predict infant outcomes in complex pregnancies with specific comorbidities.

A key strength of our study is the exclusion of pregnancies with major comorbidities and conditions that may associate with pathological placental findings, including gestational diabetes mellitus, hypertension, preeclampsia, pro-inflammatory conditions, and *in vitro* fertilization. To our knowledge, only one other study has evaluated placental histopathology in pregnancies with obesity without complications or comorbidities, and this study reported only moderate associations between increasing maternal BMI and accelerated villous maturation and chronic villitis among term pregnancies[46]. In contrast, a study by Bar et al investigating high prepregnancy BMI with maternal conditions including pre-eclampsia and gestational diabetes, but not hypertension or other pro-inflammatory conditions[47], showed increased maternal inflammatory lesions among pregnancies complicated by obesity compared to normal weight; these findings were consistent when comparing mothers with and without complications[47]. However, the Bar et al cohort did not assess underweight or preterm pregnancies, and as such, did not capture the full scope of metabolic states or gestational age effects as we did. Another larger study by Huang et al that included a cohort of women with pregnancy complications observed increased placental pathology with increasing maternal BMI, a finding that was also observed in a subset of women without obstetric complications; however, preterm and term pregnancies were not examined separately[7]. Thus, there are conflicting findings and discrepancies in cohort selection among the few studies investigating the effects of suboptimal maternal BMI on placental pathology. While future studies are required to confirm the effects of suboptimal maternal BMI alone on placental pathology, our cohort helps to address these gaps in knowledge on the impact of maternal prepregnancy BMI, preterm birth, and infection, in the absence of other major maternal comorbidities, on placental maturity and morphometry.

While suboptimal maternal BMI did not associate with either gross or microscopic placental histopathology, there is significant evidence for the effects of maternal BMI on the programming of offspring health long term[8-10]. This suggests that functional adaptations of the placenta, even in the absence of overt histomorphological changes, may still be of concern in otherwise healthy pregnancies complicated by maternal underweight, overweight, or obesity, and this warrants further investigation. For example, diet-induced obesity in mice has been shown to alter molecular pathways associated with placental development and function[48]. Further, genes associated with placental blood vessel morphogenesis have been shown to be upregulated in association with increasing maternal BMI and decreasing birthweight in humans[49]. Future studies should evaluate placental development and function at the molecular level to better understand if pregnancies with suboptimal maternal metabolic status, but that are otherwise uncomplicated, increase risk for placental dysfunction in a manner that may influence fetal development and programming of postnatal health trajectories in offspring. Such data could inform more focused efforts to mitigate these placental (mal)adaptions and would further specify the pregnancies most at risk for poor placental function and offspring programming.

Our data show that gestational age and infection most strongly associate with altered placental maturity and morphometry, and, at term and preterm, placental immaturity is more likely in pregnancies with high maternal prepregnancy BMI, despite suboptimal BMI (in the absence of other comorbidities) having few other effects on placental histopathologies. Limited changes in micro/macroscopic placental pathology do not preclude functional changes in placentae from pregnancies complicated by suboptimal maternal BMI. Our results add to the incomplete evidence on the effects of suboptimal maternal BMI, gestational age, and infection on placental maturity and morphometry in pregnancies without major comorbidities and are a step forward in understanding the mechanisms that may contribute to poor offspring outcomes in pregnancies complicated by suboptimal maternal BMI and preterm birth (with and without infection).

## Data Availability

All data produced in the present study are available upon reasonable request to the authors.

## Funding and/or Competing Interests

The authors have no competing interests to declare. This research is funded by the Faculty of Science, Carleton University. ED was supported by a Natural Sciences and Engineering Research Council Undergraduate Student Research Award. KLC is supported by grants from the Canadian Institutes of Health Research, Natural Sciences and Engineering Research Council of Canada, the Molly Towell Perinatal Research Foundation (New Investigator), and Carleton University Office of Research.

## Ethics approval

This study was approved by the Mount Sinai Hospital Research Ethics Board (17-0186-E) and the Carleton University Research Ethics Board (106932).

## Authors contributions

Conceptualisation: KC DG, EB, ED. Methodology: ED, KC, DG, EB, CBVA, AF, HS. Formal analysis and visualisations: ED, DG, CBVA, HS. Manuscript writing: ED, KC. Manuscript editing, review, and final approval: ED, KC, DG, EB, CBVA, AF, HS.

---

**Supplementary table 1.** Effect of maternal BMI group on volumetric proportions of placental histological components and placental maturity stratified by fetal sex in preterm pregnancies.

|  | UW | NW | OW | OB | P value |
| --- | --- | --- | --- | --- | --- |
| **Placental maturity (n (%))** |  |  |  |  |  |
| **Male** | (n= 6) | (n= 12) | (n= 8) | (n= 8) | 0.05 |
| Immature | 0 (0.00) | 0 (0.00) | 2 (25.0) | 2 (25.0) |  |
| Normal | 6 (100) | 6 (50.0) | 6 (75.0) | 4 (50.0) |  |
| Hypermaturation | 0 (0.00) | 6 (50.0) | 0 (0.00) | 2 (25.0) |  |
| **Female** | (n= 4) | (n= 5) | (n= 10) | (n= 4) | 0.62 |
| Immature | 1 (25.0) | 1 (20.0) | 3 (30.0) | 1 (25.0) |  |
| Normal | 1 (25.0) | 3 (60.0) | 4 (40.0) | 3 (75.0) |  |
| Hypermaturation | 2 (50.0) | 1 (20.0) | 3 (30.0) | 0 (0.0) |  |
| **Placental morphometry: volumetric proportion of histological components (%)** |  |  |  |  |  |
| **Male** | (n= 7) | (n= 11) | (n= 7) | (n= 9) |  |
| Syncytiotrophoblast | 20 (17.5, 20.5) | 21 (19, 21.8) | 20.5 (18.3, 23.8) | 20 (19.3, 25) | 0.69 |
| Cytotrophoblast | 0 (0, 0.25) | 0 (0, 0) | 0 (0, 0) | 0 (0, 0) | 0.39 |
| Connective tissue | 55.5 (54, 62) | 57 (42.3, 62.3) | 54.5 (47.3, 58.5) | 53.5 (46.3, 61) | 0.81 |
| Fetal Vascular endothelium | 22.8 ± 5.03 | 25.3 ± 11.5 | 24.3 ± 5.28 | 25.1 ± 9.85 | 0.95 |
| Syncytial knots | 1 (0, 1.25) | 0 (0, 1) | 1 (0, 2) | 0.5 (0, 1) | 0.57 |
| **Female** | (n= 3) | (n= 5) | (n= 9) | (n= 5) |  |
| Syncytiotrophoblast | 22 (20, 23) | 20 (18.5, 23.7) | 22 (19.8, 25.5) | 17 (10.8, 24.8) | 0.25 |
| Cytotrophoblast | 0 (0, 0) | 0 (0, 0.03) | 0 (0, 0) | 0 (0, 0) | 0.35 |
| Connective tissue | 57 (53, 57) | 62 (58.6, 64.5) | 57.5 (53, 59.3) | 53.5 (49.8, 55.8) | 0.12 |
| Fetal Vascular endothelium | 21.3 ± 3.81 | 17.0 ± 2.95 | 19.7 ± 2.09 | 28.8 ± 3.30 | 0.09 |
| Syncytial knots | 1 (1, 2) | 1 (0.53, 1.5) | 1 (0, 2) | 0.5 (0, 1.75) | 0.82 |

Data are presented as means ± SD (ANOVA; normal distribution/equal variance), median (IQR; Kruskal-Wallis test for non-parametric data with Steel-Dwass *post-hoc*), or n (%) (Likelihood Ratio Chi Square test) where \*p<0.05.

**Supplementary table 2.** Effect of maternal BMI group on volumetric proportions of placental histological components and placental maturity stratified by fetal sex in term pregnancies.

|  | UW | NW | OW | OB | P value |
| --- | --- | --- | --- | --- | --- |
| **Placental maturity (n (%))** |  |  |  |  |  |
| **Male** | (n= 6) | (n= 5) | (n= 2) | (n= 1) | 0.41 |
| Immature | 2 (33.3) | 1 (20.0) | 1 (50.0) | 1 (100) |  |
| Normal | 4 (66.7) | 4 (80.0) | 1 (50.0) | 0 (0.00) |  |
| Hypermaturation | 0 (0.00) | 0 (0.00) | 0 (0.00) | 0 (0.00) |  |
| **Female** | (n= 2) | (n= 2) | (n= 6) | (n= 7) | 0.31 |
| Immature | 1 (50.0) | 0 (0.00) | 4 (66.7) | 4 (57.1) |  |
| Normal | 1 (50.0) | 2 (100) | 2 (33.3) | 3 (42.9) |  |
| Hypermaturation | 0 (0.00) | 0 (0.00) | 0 (0.00) | 0 (0.00) |  |
| **Placental morphometry: volumetric proportion of histological components (%)** |  |  |  |  |  |
| **Male** | (n= 6) | (n= 5) | (n= 2) | (n= 1) |  |
| Syncytiotrophoblast | 20.5 (17.7, 23.2) | 23 (19, 23.6) | 15.5 (15, 16) | 17 (17, 17) | 0.22 |
| Cytotrophoblast | 0 (0, 0.25) | 0 (0, 0) | 0 (0, 0) | 0 (0, 0) | 0.76 |
| Connective tissue | 51.5 (44.2, 55) | 56 (50.5, 59.6) | 55.5 (55, 56) | 50 (50, 50) | 0.40 |
| Fetal Vascular endothelium | 27.6 ± 3.19 | 23.0 ± 3.49 | 29.0 ± 5.52 | 32.0 ± 7.8 | 0.62 |
| Syncytial knots | 0.5 (0, 1.25) | 0 (0, 0.8) | 0 (0, 0) | 0 (0, 0) | 0.54 |
| **Female** | (n= 2) | (n= 3) | (n= 5) | (n= 7) |  |
| Syncytiotrophoblast | 19 (18, 20) | 19 (16, 22) | 20 (15, 23) | 20 (15, 22) | 0.99 |
| Cytotrophoblast | 0 (0, 0) | 0.5 (0, 1) | 0 (0, 0.25) | 0 (0, 0) | 0.39 |
| Connective tissue | 55.5 (51, 60) | 61.5 (60, 63) | 46 (38.5, 58.8) | 56 (55.5, 56) | 0.13 |
| Fetal Vascular endothelium | 25.5 ± 4.95 | 19.0 ± 1.41 | 33.0 ± 9.63 | 24.6 ± 4.28 | 0.12 |
| Syncytial knots | 0.5 (0, 1) | 0 (0, 0) | 0 (0, 1) | 0 (0, 1.5) | 0.75 |

Data are presented as means ± SD (ANOVA; normal distribution/equal variance), median (IQR; Kruskal-Wallis test for non-parametric data with Steel-Dwass *post-hoc*), or n (%) (Likelihood Ratio Chi Square test) where \*p<0.05.

**Supplementary table 3.**
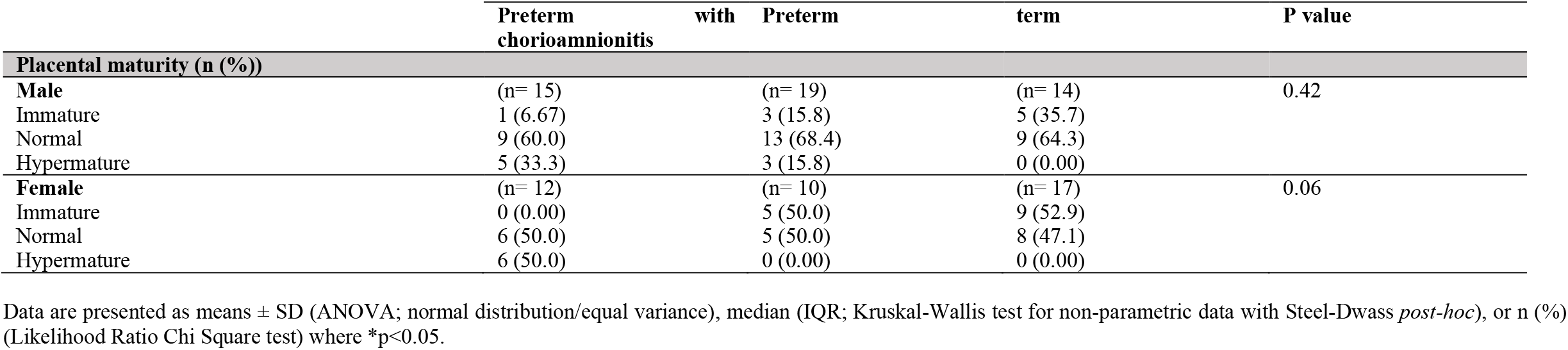
Effect of gestational age and infection inclusive of maternal BMI on placental maturity stratified by fetal sex.

**Supplementary table 4.** Placental morphometry volumetric proportions amongst preterm with chorioamnionitis, preterm, and term pregnancies stratified by fetal sex.

| Volumetric proportion of placental histological components (%) | Preterm with Chorioamnionitis | Preterm | Term | P value |
| --- | --- | --- | --- | --- |
| **Male** | (n= 16) | (n= 19) | (n= 16) |  |
| Syncytiotrophoblast | 20 (19, 22) | 20 (19, 21) | 20 (17, 23) | 0.80 |
| Cytotrophoblast | 0 (0, 0) | 0 (0, 0) | 0 (0, 0) | 0.97 |
| Connective tissue | 59 (51, 53) | 55 (46, 59) | 53 (50, 56.3) | 0.27 |
| Fetal Vascular endothelium | 22.8 ± 7.8 | 26.1 ± 9.3 | 26.4 ± 7.5 | 0.42 |
| Syncytial knots | 1 (0, 1) | 0 (0, 1) | 0 (0, 1) | 0.45 |
| **Female** | (n= 13) | (n= 12) | (n= 20) |  |
| Syncytiotrophoblast | 20.5 (19.2, 23) | 22 (19, 24.5) | 20 (15, 22) | 0.29 |
| Cytotrophoblast | 0 (0, 0) | 0 (0, 0) | 0 (0, 0) | 0.24 |
| Connective tissue | 57.5 (55.5, 62) | 55.5 (49.7, 59.2) | 56 (48, 60) | 0.17 |
| Fetal Vascular endothelium | 19.6 ± 5.06<sup>A</sup> | 22.5 ± 9.39<sup>AB</sup> | 27.3 ± 8.19<sup>B</sup> | **0.03** |
| Syncytial knots | 1 (1, 2)<sup>A</sup> | 1 (0, 1)<sup>AB</sup> | 0 (0,1)<sup>B</sup> | **0.006** |

Data are presented as means ± SD (ANOVA; normal distribution/equal variance with Tukey *post hoc*) or median (IQR; Kruskal-Wallis test for non-parametric data with Steel-Dwass *post-hoc*), where \*p<0.05.

